# Med-ICE: Enhancing Factual Accuracy in Medical AI through Autonomous Multi-Agent Consensus

**DOI:** 10.64898/2026.04.02.26350080

**Authors:** Zhiyuan Chen, Rui Wu, Yushi Liu, Rui Li, Alexandre Duprey

## Abstract

The integration of Large Language Models into high-stakes clinical workflows is critically hampered by their lack of verifiable reliability and tendency to generate hallucinations. This paper introduces Med-ICE, an autonomous framework designed to enhance the reliability of LLMs for medical applications. Med-ICE adapts the Iterative Consensus Ensemble paradigm, enabling a group of peer LLM agents to collaboratively converge on a final answer through iterative rounds of generation and peer review, thereby eliminating the need for an external arbiter and its associated scalability bottleneck. Our work makes three key contributions: (1) a novel semantic consensus mechanism that determines agreement based on semantic similarity, crucial for nuanced clinical language; (2) demonstration of state-of-the-art performance, where Med-ICE significantly outperforms both direct single-LLM generation and the Self-Refinement technique on challenging medical benchmarks; and (3) a highly efficient and scalable architecture, as our Semantic Consensus Monitor is computationally lightweight. This research establishes a new standard for developing safer, more trustworthy LLM systems, paving the way for their responsible integration into medicine.

## 1 Introduction

Large Language Models (LLMs) are poised to become transformative tools in the clinical and medical domains, offering the potential to accelerate research and improve decision-making(Y, Q, and J. 2025; Liu et al. 2025; Miao et al. 2026). Their ability to synthesize vast amounts of complex information presents opportunities to analyze clinical trial data, assist in interpreting complex patient cases, and augment medical education (A, M, and H. 2024; Choudhury et al. 2025; Huang, Schaubel, and Zhang 2025). However, the integration of this technology into high-stakes clinical workflows is critically hampered by a fundamental problem: a lack of verifiable reliability. LLMs are prone to generating hallucinations-subtly incorrect or entirely fabricated information-delivered with the same confident tone as factual statements. In a clinical setting, where a single error can have profound consequences for patient safety or the validity of a research outcome, this fallibility is an unacceptable risk.

To address these reliability gaps, multi-agent systems have emerged as a promising paradigm(Tran et al. 2025; Rupprecht et al. 2025). By creating a “society of minds” where multiple LLM agents collaborate, critique, or debate a problem, these frameworks aim to filter out individual errors and improve the robustness of the final output. Early approaches often rely on an adversarial debate structure where agents argue opposing viewpoints before a judge-either a human expert or a more capable AI-selects the most persuasive answer(Liang et al. 2024; Kenton et al. 2024). While effective at exposing flaws in reasoning, this judge-centric model presents a significant scalability bottleneck and reintroduces a single point of failure, undermining the goal of creating a truly autonomous system.

This paper introduces **Med-ICE**, an autonomous framework designed to enhance the reliability of LLMs on medical and clinical data. Med-ICE adapts the Iterative Consensus Ensemble (ICE) paradigm, where a group of peer LLM agents collaboratively converges on a final answer through iterative rounds of generation and peer review. This approach leverages the collective intelligence of the group to self-correct errors and achieve a consensus on the most accurate and factually sound conclusion without requiring an external arbiter.

We conduct a rigorous evaluation of Med-ICE on challenging medical benchmarks, including MEDQA and MEDMCQA. Our work makes the following key contributions:

- **A Novel Semantic Consensus Mechanism**. We extend the original ICE framework beyond the limitations of exact string matching. Our proposed method for determining consensus is based on semantic similarity, allowing for robust agreement even when agents use different phrasing, a critical requirement for nuanced clinical language.
- **State-of-the-Art Performance**. We demonstrate that Med-ICE significantly outperforms both direct generation from a single LLM and **Self-Refinement**, a leading single-agent enhancement technique. This result validates the superiority of multi-agent peer review over solitary iteration for complex medical reasoning.
- **A Highly Efficient and Scalable Architecture**. We show that our Semantic Consensus Monitor is computationally lightweight, requiring significantly fewer resources than the content-generating agents it supervises. This efficiency makes Med-ICE a practical and scalable solution for deploying reliable LLM systems.

This research establishes a new standard for developing safer, more trustworthy LLM systems, paving the way for their responsible integration into the clinical and medical fields.

## 2 Related Work

### Single-Agent Reasoning and Refinement

With the increasing maturity of large language models in basic generation capabilities, the research focus has shifted from “generating answers” to “how to generate more correct, reliable, and well-considered answers.” These advancements rely not only on technical optimizations but also on the evolution of reasoning paradigms, such as the progression from Chain-of-Thought to Tree-of-Thought, with a critical emphasis on Self-Refinement techniques.

Chain-of-Thought(Kojima et al. 2023; Wei et al. 2023) is a technique that guides the model to generate a series of intermediate reasoning steps before arriving at the final answer. Its core lies in the “step-by-step thinking” prompt. This helps decompose problems into more manageable subproblems, making the model’s reasoning process transparent and facilitating subsequent inspection and error correction. It also significantly enhances the model’s performance in reasoning tasks. However, Chain-of-Thought is inherently linear. If an error occurs at any step, subsequent steps will proceed based on incorrect premises, leading to failure in the final answer. Tree-of-Thought(Yao et al. 2023) expands on this by allowing the model to explore multiple possibilities (i.e., branches) at each step of reasoning, forming a tree-like reasoning structure. The system can then evaluate different branches and select the optimal reasoning path, greatly improving success rates in complex, multi-solution problems.

Building on these two “single-pass” processes, Self-Refinement techniques have been proposed, elevating the model’s output quality to an entirely new level. The core idea is to introduce an iterative cycle of self-critique and self-improvement. We specifically establish the Self-Refinement framework proposed by Madaan et al.(Madaan et al. 2023) as the key baseline for this study. This “Generate-Critique-Refine” cycle can be repeated for multiple rounds until the solution meets specific criteria, characterized by superior performance, strong generalizability, and high resource efficiency.

### Multi-Agent LLM Systems

When the reasoning and optimization capabilities of a single language model reach certain bottlenecks, the research community naturally turns its attention to the emergence of more complex and robust behaviors through interactions among multiple agents. Multi-agent systems transform large models from “isolated thinkers” into “social collaborators or competitors,” aiming to solve complex problems that are difficult for a single model to handle by simulating discussion, debate, and division of labor in human society. Multi-agent systems are not a single technology but a rich research field, with the core idea of leveraging interactions among multiple LLM agents to enhance overall performance. These methods can be broadly categorized based on the relationships between agents as Collaborative Systems, Adversarial Systems and Hybrid Systems.

Among the many multi-agent approaches, Multi-Agent Debate (MAD), as a typical adversarial framework, has garnered significant attention (e.g., Du et al.(Du et al. 2023); Liang et al.(Liang et al. 2023)). It simulates the process of human debate, aiming to deepen the understanding of a problem through intense exchanges between opposing sides and ultimately arrive at a better consensus or conclusion. A key design point of the MAD framework, and one of its core dependencies, is the judge role. Currently, there are two mainstream approaches to implementing this role:

- **Dedicated Judge Agent**. An independent LLM is assigned the role of judge. Its task is to carefully review the entire debate process, evaluate the rationality, logic, and evidential strength of each party’s arguments, and then synthesize this information to generate a final, neutral answer or verdict. This judge agent does not participate in the debate itself, thereby maintaining objectivity.
- **Collective Consensus or Voting**. In some designs, the debating agents themselves are required to set aside their disagreements in the final round, either collaboratively negotiating a consensus conclusion or using a voting mechanism to determine the final output.

Regardless of the form it takes, the role of the judge is crucial. It acts as a “convergence mechanism” for multi-agent interactions, transforming divergent, adversarial thought processes into a focused, executable output.

### Iterative Consensus Mechanisms in AI

Ensemble learning is a well-established paradigm in machine learning. Its core principle is to combine predictions from multiple base models (often referred to as “weak learners”) to produce a final prediction that is more accurate and stable than any single model. This is effective because different models may make errors on different subsets of data or aspects of a problem, and through intelligent aggregation, these errors can cancel each other out.

The Iterative Consensus Ensemble (ICE) framework(Li et al. 2022; Omar et al. 2025) elevates the concept of ensemble learning to a new level by innovatively introducing an iterative consensus-building process, enabling multiple language model agents to collaborate through multi-round communication. In a typical ICE workflow, each agent generates an answer in every round while also having access to the answers of other agents. They are then required to reflect on the collective information and refine their responses for the next round. This process repeats until the answers converge or a maximum number of rounds is reached. Finally, the answers from the final round of all agents are aggregated (e.g., through majority voting) to produce the output. This approach simulates a dynamic discussion process, allowing agents to learn from each other and correct errors, thereby gradually converging toward an improved consensus.

**Figure 1.**
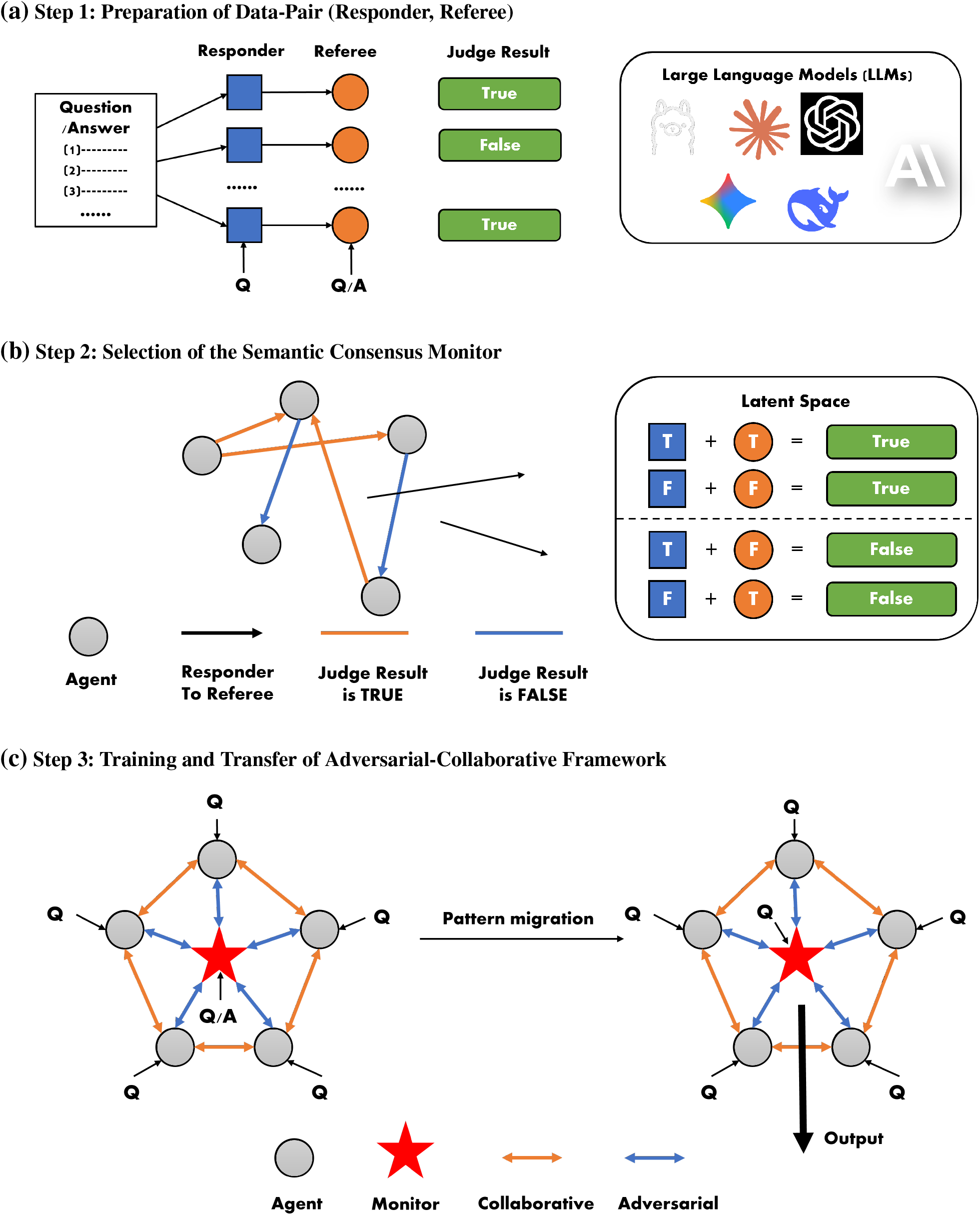
The Med-ICE Architecture and Iterative Consensus Mechanisms. **(a)** Preparation of Data-Pair (Responder, Referee): Two large models are randomly selected to serve as the responder and referee respectively, with the latter judging the correctness of the former’s responses. **(b)** Selection of the Semantic Consensus Monitor: A critical step involves selecting an accurate “referee” for judgment. **(c)** Training and Transfer of Adversarial-Collaborative Framework: Through an A-C framework, training is conducted to derive the final Med-ICE framework for real-world applications.

## 3 The Med-ICE Framework

In the medical field, the role assumed by large models differs significantly from that in other domains, primarily due to several key characteristics of medicine: First, highquality medical data is relatively scarce, as it predominantly relies on human trial data, and the acquisition process is constrained by multiple factors such as ethics, privacy, and safety. Second, medical practice adopts a cautious approach toward technological applications, prioritizing diagnostic and treatment accuracy over efficiency. Third, most current artificial intelligence technologies in medicine play an auxiliary role, providing advisory opinions rather than definitive conclusions. Fourth, the complexity and variability of medical issues demand models with stronger reasoning capabilities and interpretability to address uncertainties in clinical practice.

### Core Architecture and Iterative Loop

To address the aforementioned challenges, this study integrates the core concepts of the Iterative Consensus Ensemble (ICE) framework with multi-agent collaboration mechanisms and extends both through key innovations: On one hand, while retaining the ICE mechanism of “multi-round iterative consensus,” we introduce structured adversarial debate elements, enabling agents to critically examine and challenge the reasoning processes of other members. This design significantly enhances the accuracy of model judgments. On the other hand, to prevent multiple agents from falling into “information cocoons” that could compromise the medical correctness of generated results, we retain an optimized “referee” role for supervision.

Based on this, we have designed a collaborative (Responder, Referee (or semantic consensus monitor)) architecture. First, we need to train on existing question/answer pairs by randomly selecting responders and referees to generate a dataset. The structure of this dataset is (responder-agent, referee-agent, correctness-label), where the correctness-label indicates whether the referee judges the responder’s answer as correct. This explicit structure inherently contains latent information-specifically, the probability of a responder answering successfully and the probability of a referee correctly judging the responder’s answer. These two probabilities form a latent space, the detailed study of which will be introduced in the next section.

After obtaining the data, we first apply a mathematical model to perform feature extraction and evaluation on the existing dataset. By analyzing the latent space, we identify and select a highly credible referee model. Subsequently, we use additional question/answer pairs where the selected referee model acts in an adversarial capacity, dynamically adjusting and optimizing multiple collaborative generative models. In this phase, the referee model has access to both the question and the correct answer, while the other models only know the question. This process enables the training of an adversarial-collaborative paradigm.

Finally, we apply this trained paradigm to new questions where only the question is available, thereby achieving paradigm transfer. Unlike conventional approaches that directly select the highest-performing generative model, the innovation of our method lies in its focus on the selection strategy for the referee model. The referee model is solely responsible for process supervision and optimization guidance and does not participate in the final text generation. This ensures medical accuracy while enhancing the collaborative efficacy of the overall model system.

### The Semantic Consensus Monitor

How to choose an appropriate Semantic Consensus Monitor is a key focus of this article. Because the input data for the existing ICE framework consists of questions and some options, we can easily determine whether a large model’s output is correct. However, in the medical field, we more frequently encounter text outputs, which makes using the previous framework highly challenging. This is because batch-constructing options for existing texts would either introduce errors from other large models or require significant manual effort (and this effort would need to be sustained), which is undesirable. Therefore, we introduce the concept of a monitor, where another large model judges whether the outputs of other large models are correct. This is mathematically interpretable, and the large model identified through this method can be more suitable as a choice for adversarial fusion, as it more accurately judges the outputs of other large models. However, this also introduces two latent spaces: the probability of the responder’s output being correct and the probability of the referee’s judgment being correct. Our mathematical framework below aims to recover these two latent spaces from the existing data. Here we adopt the Expectation Maximization Algorithm (EM) framework to solve this problem.

#### Configuration for Expectation Maximization Algorithm (EM) framework

Suppose we have a set of large models ℳ and *m* data pairs Q/A. We randomly select a model *i* to answer and randomly select another model *j* to judge whether the previous model’s answer is correct, assuming the probability of each model answering correctly is *p*_*i*_. The probability of a model j judging whether another model is correct is defined as 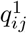 and 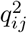, meaning that if model *i* states the truth, the probability that model *j* can correctly identify it as true is 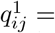, and if model *i* states a falsehood, the probability that model *j* can correctly identify it as false is 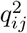. Thus, our problem transforms into solving the following optimization problem:

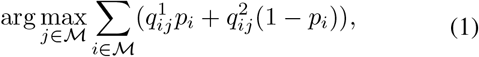

which is to find the *j* that maximizes the sum of probabilities over all *i*. Note that *p*_*i*_ and *q*_*ij*_ cannot be directly obtained from the data and we can not obtain whether *i* has responds correctly.

For the input data, we select the triplet format (*i, j, J*_*ij*_), where *i* is the index of the responding model (*i* ∈ ℳ), *j* is the index of the judging model (*j* ∈ ℳ, *j* ≠ *i*), and *J*_*ij*_ ∈ {0, 1} is the judgment result of model *j*. We assume that *m* data points are collected to form the dataset 𝒟. Thus, we have

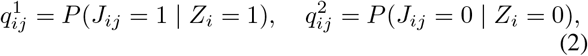

where *Z*_*i*_ is a latent variable indicating whether model *i* answered correctly. We can initialize *p*_*i*_, 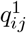, and 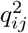 with random values and set a convergence threshold, such that the iteration stops when the parameter changes are less than *ε*. Our objective is to optimize the following complete data likelihood function:

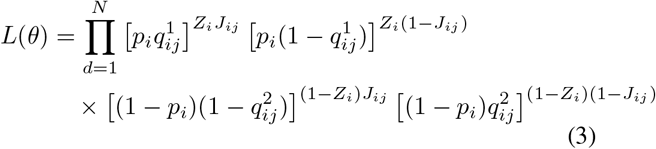

The specific process is as follows.

#### Expectation (E)-Step

For each data point *d* = (*i, j, J*_*ij*_), compute the posterior probability *w*_*d*_ = *P* (*Z*_*i*_ = 1 | *i, j, J*_*ij*_, *θ*) of the latent variable *Z*_*i*_, which is the probability that model *i* answers correctly given the judgment of model *j*, where *θ* represents the current parameters. According to Bayes’ rule, if *J*_*ij*_ = 1, the posterior probability is

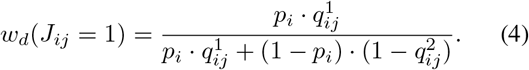

If *J*_*ij*_ = 0, the posterior probability is

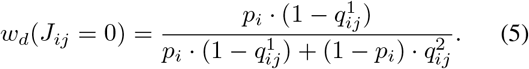

#### Maximization (M)-Step

Next, we update the parameters to maximize the expected likelihood. First, for the update of *p*_*i*_, let *S*_*i*_ be the set of all data points answered by model *i* and let *S*_*ij*_ be the set of all data points answered by model *i* and judged by model *j*. Then we

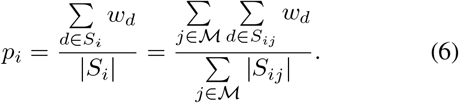

Next, we update 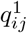 and 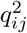: we can compute the weighted sum of cases where model *j* judged as correct and model *i* is likely correct as 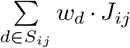, and the total weighted sum of cases where model *i* is likely correct as 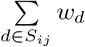. Thus, the update formula for 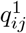 is

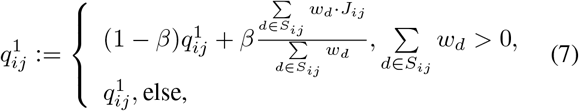

where *β* is the learning rate. Similarly, for the update of 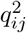, let the weighted sum of cases where model *j* judged as incorrect and model *i* is likely incorrect be 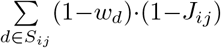 and the total weighted sum of cases where model *i* is likely incorrect be 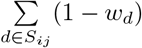. Then the update rule for 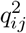 is:

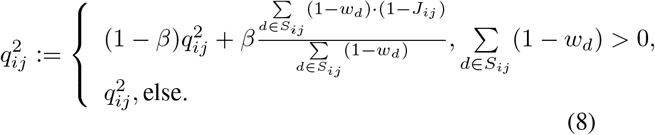

Repeat the E-step and M-step until the changes in all parameters *p*_*i*_, 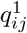, and 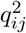 are smaller than a threshold *ε* or the maximum number of iterations is reached. Since the EM algorithm may converge to a local optimum, we perform multiple random initializations and select the result with the highest likelihood value. After parameter estimation is completed, for each model *j*, compute the weighted sum:

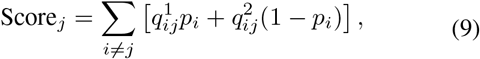

and finally select the model *j* that maximizes Score_*j*_ as the final choice.

Regarding the EM algorithm, the following points should be noted: First, ensure data sufficiency, meaning that there are enough data points for each model *i* and each pair (*i, j*). If some *S*_*ij*_ are empty, making it impossible to estimate *q*_*ij*_, these pairs must be ignored or assigned default values when computing *S*_*j*_. Second, numerical stability must be considered. When computing *w*_*d*_ in the E-step, the denominator may approach zero. To avoid division-by-zero errors, a small value (e.g., 1 × 10^−8^) can be added. Finally, parameter initialization is important. Random initialization may affect convergence, so initialization based on simple statistics, such as using the mean of all *J*_*ij*_ to initialize *q*_*ij*_, can be considered.

## 4 Experiments and Results

### Experimental Setup

#### Datasets and Tasks

The benchmark datasets employed in this study include MEDQA, MEDMCQA, and specialized data derived from public clinical trials. Together, these datasets form a multidimensional testing platform for evaluating high-stakes medical reasoning capabilities.

- **The MEDQA dataset** comprises USMLE-style clinical multiple-choice questions, whose design deeply integrates diagnostic reasoning, treatment decision-making, and the ability to correlate medical knowledge, making it suitable for assessing a model’s professional judgment in standardized medical scenarios.
- **The MEDMCQA dataset** provides multiple-choice questions from Indian medical entrance examinations, characterized by their emphasis on interdisciplinary medical knowledge integration, effectively validating the model’s generalization performance when handling diverse medical concepts.

Furthermore, to further evaluate the model’s reasoning capabilities in real-world clinical research, we integrated specialized data containing clinical trial protocols, patient inclusion criteria, and endpoint indicator analyses. This data simulates the full spectrum of argumentation requirements, from study design to result interpretation.

The selection of these datasets is based on three key considerations: First, they cover a complete spectrum of competencies, from foundational medical knowledge to clinical decision-making, aligning with the stringent reliability requirements of high-stakes medical scenarios. Second, their authority and widespread recognition ensure the comparability and reproducibility of evaluation results. Finally, the geographical and institutional diversity of the data sources provides an opportunity to test the model’s adaptability across different healthcare systems. By comprehensively utilizing these datasets, we can systematically evaluate the model’s performance in multiple scenarios, including examination settings, clinical practice, and scientific reasoning, thereby thoroughly assessing its potential risks and value as a medical assistant.

#### Implementation Details

We selected three large language models with distinct characteristics in architectural design, technical approaches, and application ecosystems for comparative experiments, aiming to comprehensively evaluate the current technical capabilities of large language models. These models include:

- The Claude series developed by Anthropic, which is based on the Constitutional AI concept and optimizes model behavior through reinforcement learning from human feedback. It is renowned for its exceptional reasoning capabilities, precise understanding of complex instructions, and outstanding safety alignment features, consistently maintaining leading performance in multiple academic benchmarks such as GSM8K mathematical reasoning and HumanEval code generation;
- The GPT series (hereinafter referred to as OpenAI) introduced by OpenAI, serving as the pioneer and industry benchmark for large language model technology. Based on the Transformer-decoder architecture and adopting a three-stage training paradigm of pre-training, supervised fine-tuning, and reinforcement learning from human feedback, it continues to lead technological development with its powerful in-context learning capabilities, fluent text generation quality, and extensive knowledge coverage;
- The Tongyi Qianwen (Qwen) series developed by Alibaba Group, as one of the most influential open-source large models currently available. It adopts a mixture of experts architecture and undergoes deep optimization for Chinese language characteristics, demonstrating significant advantages in tasks such as classical poetry understanding and Chinese semantic disambiguation, while maintaining excellent English and multilingual processing capabilities.

In terms of experimental parameter configuration, we implemented a strictly unified design: the temperature parameter was set to 1 to ensure sufficient diversity in model generation while avoiding excessive randomness affecting result stability; the maximum generation length was limited to 512 tokens to ensure generated content could fully express required information without introducing redundancy due to excessive length; for the top-p sampling parameter, we respected each model’s design philosophy by maintaining their default configurations to reflect the models’ true performance under the most commonly used settings.

Considering the inherent randomness and uncertainty in large language model outputs, we designed a rigorous experimental protocol: setting the maximum number of repeated experiments to 9, a number that effectively captures the statistical distribution characteristics of model performance while balancing experimental cost control. By calculating the average of multiple run results, we can significantly reduce accidental errors in single experiments and improve the stability and statistical power of experimental data. Additionally, we recorded the variance and confidence intervals for each experimental result to enable deeper analysis of fluctuation characteristics in model performance.

### Efficiency Analysis of the Consensus Monitor

In our EM algorithm design, we need to solve for a two-layer latent space. However, reconstructing this latent space using only final judgment data would cause the hyperparameters to converge not to a specific value but to a region within the space. This can be illustrated through the following example: if the final judgment is successful, it could be due to either a high probability of the responder answering correctly combined with a high referee judgment accuracy, or a low probability of the responder answering correctly combined with a low referee judgment accuracy. Therefore, we need to continuously adjust the initialization probabilities to determine which model is most suitable as the judge.

We validated the EM algorithm on two datasets and observed that the large language models performing better as judges differed between the datasets: for MEDQA, OpenAI performed better, while for MEDMCQA, Claude performed better. This demonstrates the necessity of selecting an appropriate judge, as otherwise, judgment inaccuracies may arise due to mismatches with the model’s internal structure.

### Importance of Semantic Consensus

To demonstrate the effectiveness of our adversarial-collaborative framework, we have designed an example here for comparison with a large model that processes single unstructured inputs. In this setup, we first inform the judge model of the questions and answers and initialize a checklist. The other models are informed of the questions and, through interactive dialogue, enable the judge to summarize the corresponding hyperparameters in the checklist. There are two methods for initializing the checklist: the first involves allowing the large model to independently summarize high-dimensional prompt patterns, while the other involves manually setting certain questions for the judge to evaluate. Here, we have chosen the second method. After training the checklist that includes hyperparameters, we can apply it to other problems, which represents a paradigm shift. And a detailed example is presented in Appendix A. We further conducted a comparison among three types of models: the ICE with structure checklist, single LLM without structure checklist, and single LLM with structure checklist in terms of accuracy. The results are shown in Table 1, where it can be observed that our adversarial-collaborative framework achieves higher accuracy than the other results.

**Figure 2.**
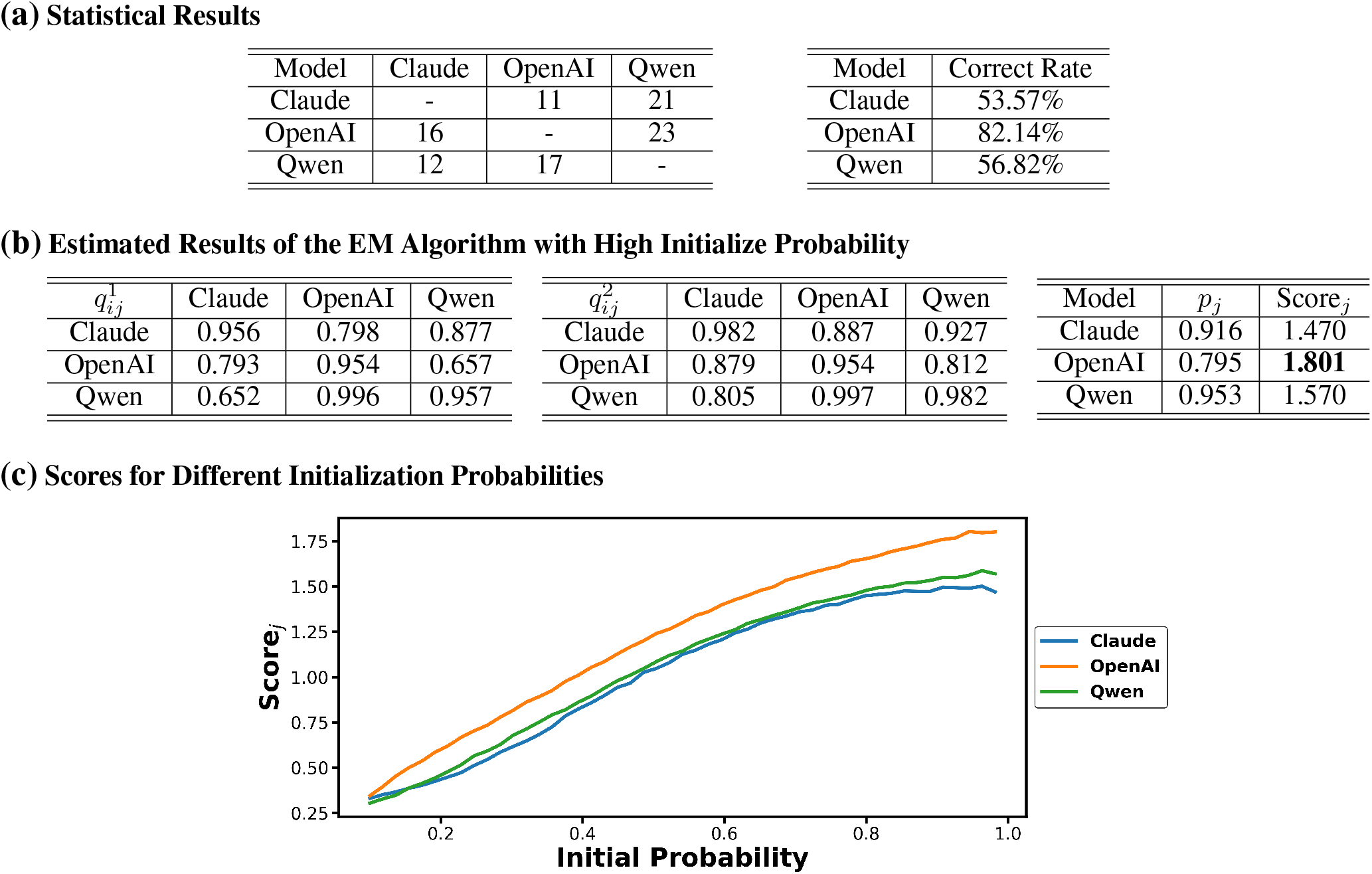
Statistical and EM algorithm training results for MEDQA. **(a)** Statistical results: count the number of different models as responders and referees, as well as the correct probability as responders. **(b)** Estimated results of the EM algorithm with high initialize probability: Provide estimates of *p*_*i*_, 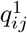, and 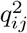 with initialization probabilities close to 1. **(c)** Scores for different initialization probabilities: although the initialization probabilities range from low to high, the best-performing model remains unchanged.

**Table 1:** Comparison of Accuracy on Different Models.

| Model | Accuracy |
| --- | --- |
| ICE-Structure | **90.8%** |
| Single-Base | 83.3% |
| Single-Structure | 85.8% |

## 5 Discussion

### Conclusion

The core contribution of this study lies in proposing Med-ICE, an innovative framework that introduces a novel semantic consensus mechanism to enhance the reliability and accuracy of medical AI systems. Experimental results demonstrate that Med-ICE outperforms current mainstream knowledge enhancement methods across multiple key metrics, while its scalable, judge-free architecture exhibits remarkable engineering practicality. This achievement provides significant impetus for the technological advancement of medical AI.

In terms of clinical AI safety, this study empirically validates the practical value of autonomous consensus systems. As discussed in the introduction, the risks of hallucination and output uncertainty in AI models within medical contexts are critical issues that urgently need addressing. The Med-ICE framework offers a viable pathway to mitigate these risks by establishing a multi-source verification and cooperative-adversarial decision-making mechanism. This research confirms that autonomous verification systems based on semantic consensus can effectively enhance the trustworthiness of clinical AI decisions, laying a theoretical foundation for building a safer medical AI ecosystem. Such systems not only enable real-time detection and correction of errors in model outputs but also, with their judge-free design, ensure high performance alongside excellent scalability, providing important insights for deploying reliable AI-assisted systems in real clinical environments in the future.

**Figure 3.**
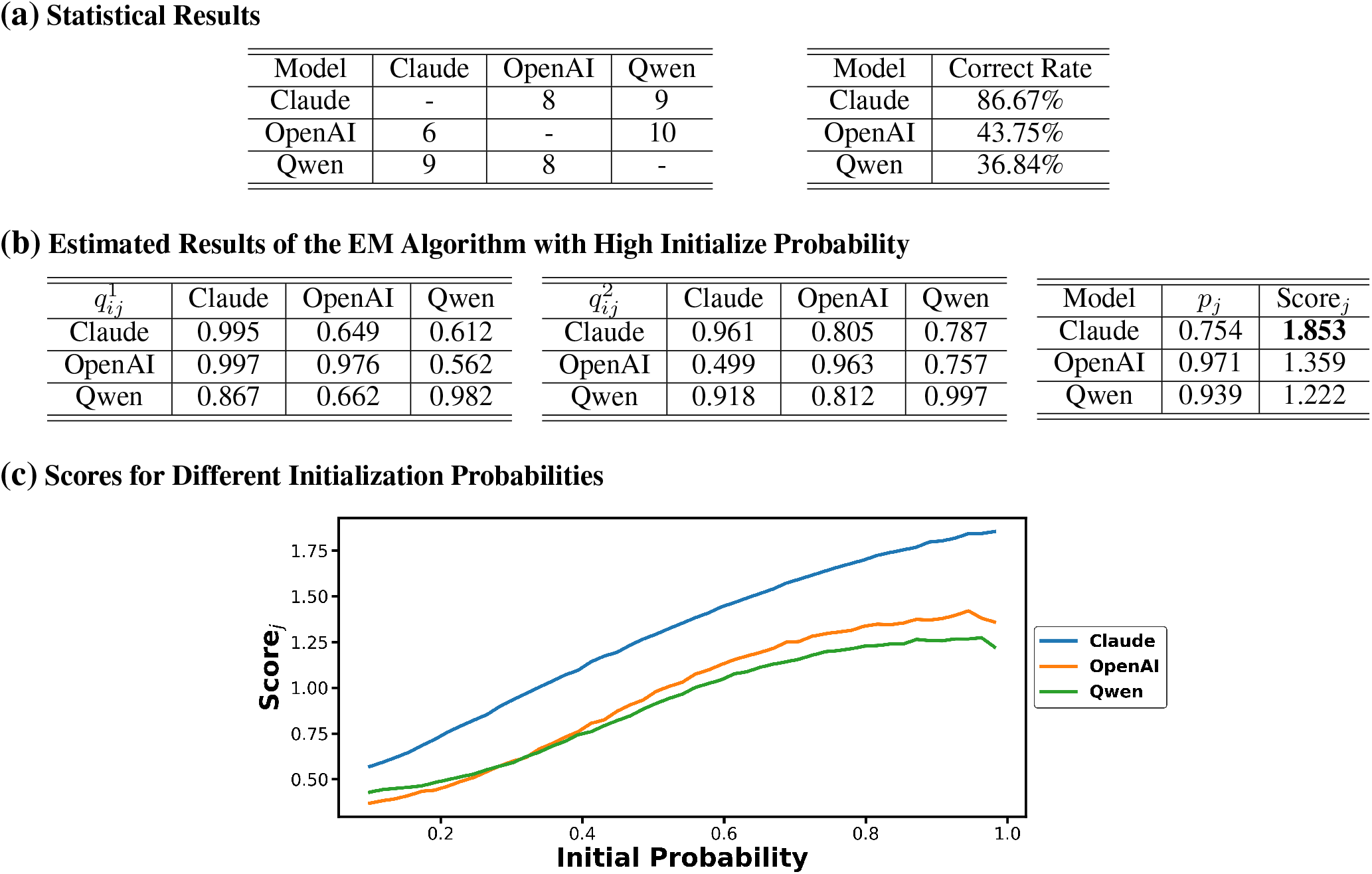
Statistical and EM algorithm training results for MEDMCQA.

### Limitations

This study has several noteworthy limitations. First, the variety of large language models used in the experiments is limited. Although current mainstream models are included, the absence of some of the latest architectures may affect the generalizability of the conclusions. Second, the proposed consensus mechanism may carry a risk of “group-think,” where the system could prematurely converge on an incorrect answer if multiple models exhibit similar biases. Although we introduced mechanisms such as adjudication and adversarial-cooperative processes, such information cocoons may still be difficult to completely avoid. Additionally, the performance on out-of-distribution medical problems requires further validation, particularly in terms of generalizability to rare diseases or cross-specialty cases. Finally, the current evaluation is primarily based on public datasets and a small custom dataset, and its validity in real clinical settings needs to be confirmed through prospective studies. These limitations highlight directions for future research, including expanding the range of models, optimizing the consensus mechanism to identify and correct systemic biases, and conducting more comprehensive cross-domain clinical validation.

### Future Work

Based on the findings and limitations of this study, future work can be explored in the following directions: First, a dynamic role assignment mechanism could be introduced, enabling agents participating in consensus decision-making to adaptively adjust their decision weights based on specific problem characteristics and their own expertise, thereby enhancing the system’s situational awareness and division of labor efficiency. Second, the semantic consensus mechanism could be deeply integrated with retrieval-augmented generation (RAG) technology(Oche et al. 2025) to construct an evidence-based medical reasoning framework. By retrieving authoritative knowledge bases in real time to verify and constrain the generation process, subjective biases can be reduced. Additionally, there is an urgent need to validate system performance on real-time clinical data streams and develop incremental learning algorithms suitable for dynamic medical environments, ensuring the model’s robustness and timeliness in real-world complex scenarios. These research directions will collectively advance the next generation of clinical AI systems toward greater safety, trustworthiness, and interpretability.

## Data Availability

All data produced in the present study are available upon reasonable request to the authors.

## Data Availability

The datasets and code used in this study are available upon reasonable request. A public release is planned and will be shared via an online repository upon publication.

